# Inflammatory microenvironment impairs the therapeutic effect of daratumumab-lenalidomide in multiple myeloma

**DOI:** 10.1101/2025.04.23.25325569

**Authors:** Wenwen Cheng, Silvia Gaggero, Lama Hasan Bou Issa, Aïcha Ouelkdite, Guillaume Escure, Adeline Cozzani, Yanick Njosse Tchantchou, Noémie Carlier, Emilie Wauquier, Gauthier Decool, Rémi Tilmont, Benjamin Podvin, Malo Leprohon, Léa Fléchon, Angélina Kasprowicz, Sophie Groux-Degroote, Jérôme Moreaux, Bertrand Arnulf, Jill Corre, Céline Villenet, Ludovic Martinet, Xavier Leleu, Martin Figeac, Thierry Facon, Wenfei Jin, Suman Mitra, Salomon Manier

**Author notes:** These authors participated equally (Wenwen Cheng, Silvia Gaggero). Co-senior authors (Wenfei Jin, Suman Mitra, Salomon Manier).

## Abstract

Daratumumab and lenalidomide (DR) combination therapy improves multiple myeloma outcomes, yet some patients respond poorly. We examined 56 DR pre-treatment samples from the IFM2017 Phase 3 trial using multiomics approaches to identify determinants of response. Analysis of bone marrow environment revealed critical synergy between inflamed myeloma cells and inflammatory immune microenvironment contributing to treatment failure. We identified non-responders exhibiting inflammatory signatures with impaired monocyte phagocytosis, upregulated NF-kB signaling in monocytes, increased pro-inflammatory ISG+ T cells, and attenuated NK cell CD16 expression—undermining antibody-dependent cellular cytotoxicity essential for daratumumab efficacy. Concurrently, myeloma cells displayed enhanced NF-kB activation, creating an inflammatory circuit. We developed a model based on 34 NF-kB-related genes (AUC=0.96) discriminating responders from non-responders. A refined eight-gene signature was robustly validated using bulk RNA sequencing in bone marrow and peripheral blood cohorts (AUCs=0.92 and 0.77, respectively). These findings suggest anti-inflammatory strategies may enhance treatment efficacy.

## Introduction

The advent of anti-CD38 monoclonal antibodies has transformed the treatment landscape of multiple myeloma (MM). Among these, the combination of daratumumab and lenalidomide (DR) has demonstrated remarkable efficacy, becoming standard treatment for newly diagnosed MM patients who are not eligible for transplant(1–3). However, despite the substantial success of this regimen, some patients fail to achieve deep responses, and all eventually relapse. To enhance outcomes, a deeper understanding of the immune microenvironment necessary for an optimal response to DR therapy is critical.

Daratumumab, a monoclonal antibody targeting CD38, exerts its effects through multiple mechanisms, including antibody-dependent cell-mediated cytotoxicity (ADCC), antibody-dependent cellular phagocytosis (ADCP), and complement-dependent cytotoxicity (CDC)(4,5). Additionally, daratumumab has been shown to deplete CD38^+^ regulatory T cells, potentially enhancing the activity of helper and cytotoxic T-cells(6). Lenalidomide, an immunomodulatory drug, acts directly on plasma cells by promoting the degradation of the lymphoid transcription factors IKZF1 and IKZF3 via the CRBN-CRL4 ubiquitin ligase complex(7,8). These transcription factors are crucial for plasma cell differentiation and MM cells progression. Lenalidomide also exerts an indirect immunomodulatory effect by enhancing the immune system, including the induction of interleukin-2 production in T cells(8,9). The synergistic combination of DR activates natural killer (NK) cells and expands effector memory T cells(9).

In MM, the bone marrow immune microenvironment undergoes significant alterations throughout disease progression(10–12). For instance, an increase in NK cells is often observed in the early stages, accompanied by altered chemokine receptor expression(12). In MM patients, the frequency of mature cytotoxic CD56^dim^ NK cells decreases, while the frequency of CD16/CD226^low^ NK cells increase — this subset exhibits adhesion defects and impaired effector functions(13). At the smoldering MM stage, the loss of granzyme K+ memory cytotoxic T cells and the dysregulation of CD14^+^ monocytes contribute to tumor progression(12). These immune alterations may be driven by inflammatory mesenchymal stromal cells, which co-localize with tumor and immune cells and express genes involved in tumor survival and immune modulation(14).

The implications of these immune alterations in the context of MM treatment remain poorly understood, and their impact on patient outcomes is unclear. To address these gaps, we conducted single-cell sequencing (scRNA-seq) of 21 bone marrow samples from patients treated with DR as part of the IFM2017-03 clinical trial (NCT02252172). Our findings reveal an enrichment of inflammatory signatures in the bone marrow of poor responders, leading to dysfunctional CD56^dim^ NK cells and impaired phagocytic capacity of CD14^+^ monocytes.

## Results

### Study population

To assess the impact of the immune microenvironment on MM treatment, we performed a scRNA-seq on bone marrow samples from 21 patients with newly diagnosed MM (NDMM) (**Fig. 1a, Supp.Table 1**). All patients were enrolled in the IFM2017-03 clinical trial and treated by a combination of DR. This phase 3 study evaluated a dexamethasone-sparing strategy for frail patients with NDMM. In arm B, patients received DR until progression and only two initial cycles of dexamethasone. The main clinical characteristics of the patients are summarized in Supplementary Table 1. Based on treatment response, patients were divided into two groups: 11 complete responders (CR) and 10 non-responders or poor responders (NR).

**Figure 1.**
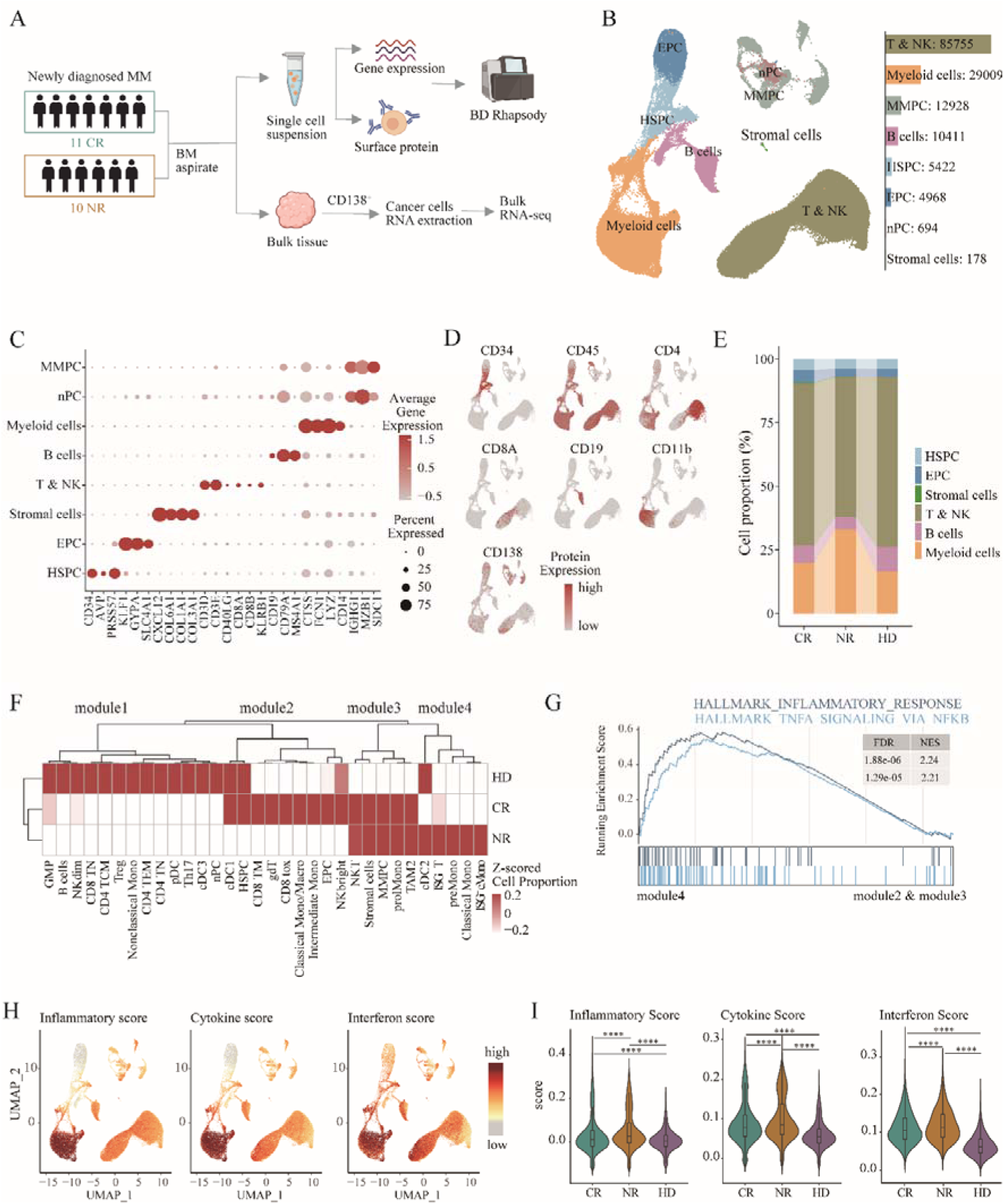
Single cell transcriptomic composition of human bone marrow cells. (a) Workflow of single-cell and bulk sequencing for 21 multiple myeloma (MM) samples. (b) Single-cell atlas of bone marrow cells from MM patients pre-treatment and controls. T and myeloid cells make up the largest proportions, with plasma cells categorized into multiple myeloma plasma cells (MMPC) and normal plasma cells (nPC). (c) Dot plot corresponding to UMAP regions highlighting the expression of cluster-specific genes. The dot size represents the percentage of cells expressing each gene, and the color indicates the average expression level within each cluster. (d) Feature plots show surface protein expression, with darker colors indicating higher protein levels. (e) Proportions of each cell cluster across different sample groups. (f) Heatmap showing cluster proportions based on Euclidean distance, with cells divided into four modules. Module 4 is specific to the NR group. (g) GSEA analysis identifies pathways significantly enriched in Module 4. (h-i) Feature and violin plots display inflammatory, cytokine, and interferon scores. Statistical significance was determined using the Wilcoxon test.

### Inflammatory bone marrow at diagnosis is associated with poor responses

A total of 153,913 single cells were sequenced by paired transcriptome and a panel of 35 antibodies, with a median of 6505 cells (range 3646-11360) per sample. Bone marrow scRNA-seq data in 20 healthy donors (HD) from Oetjen *et al.* (15)were selected as control group (**Supp. Table 2**). After quality control and normalization, a total of 149,365 cells from MM patients and HD were analyzed, encompassing all major cell types of the bone marrow cells: hematopoietic stem cell (HSC), erythroid progenitor cells (EPC), stromal cells, T & NK, B cells, myeloid cells, normal plasma cells (nPC) and MM plasma cells (MMPC) (**Fig. 1b and Supp. Fig. 1a**).

Matched gene expression levels and antibody expression level analysis effectively identified key clusters, including *CD34* for HSPC, *PTPRC* and *KLRB1* for T & NK, *CD19* and *CD79A* for highly expressed in B cells, *FCN1* and *ITGAM* for myeloid cells, normal PC in 20 HD clustered together had high expression of *MZB1* and low expression of *SDC1*, while malignant plasma cells from MM patient had high expression of *SDC1* (referred to as MMPC) (**Fig. 1c-d and Supp. Fig. 1b-c**). There was no difference in proportion of B, T, NK, monocytes between CR and NR (**Fig. 1e**). However, when compared to healthy donors, MM patients had higher proportion of myeloid cells and the enrichment of T and NK cell ratios, consistent with previous findings (16) (**Fig. 1e**).

To explore factors influencing responses to DR, we compared the characteristics of different response groups. By clustering based on the proportions of subsets across the groups, we observed that Module 4, which was highly enriched in the NR group, showed significant enrichment for the *hallmark_inflammatory_response* and *hallmark_TNFa_signaling_via_NFkB* pathways (**Fig. 1f-g**). Furthermore, we compared three inflammatory cytokine and interferon signature scores among CR and NR groups at the bone marrow microenvironment (BME) level. These signatures were predominantly enriched in myeloid cells and subsets of T&NK cells (**Fig. 1h**). A clear distinction emerged, with significantly higher scores for all three signatures – inflammation, cytokine, and interferon - in NR patients compared to the CR patients (**Fig. 1i**). This suggests that an inflamed microenvironment, characterized by elevated inflammatory cytokines and interferons, may play a role in mediating resistance to DR therapy.

### Inflammatory-associated classical monocytes have impaired phagocytosis capacity in NR

We next focused on the subsets of myeloid cells associated with inflammatory profiles and their capacity to activate NF-kB signaling. Myeloid cells from 21 patients and 20 HD were integrated and annotated into 13 clusters (**Fig. 2a**). Classical monocytes with the largest number of cells were characterized by high expression of *FCN1* and *S100A12*, indicative of their origin from blood-derived monocytes. Intermediate monocytes had relatively weak expression of *CD14* and *CD68*, while classical monocyte/macrophages highly express *CD14* and macrophage marker *CD163* (**Fig. 2b and Supp. Fig. 2a-b**). Nonclassical monocytes exhibited high expression of *FCGR3A* but lack *CD14* (**Fig. 2b).**

**Figure 2.**
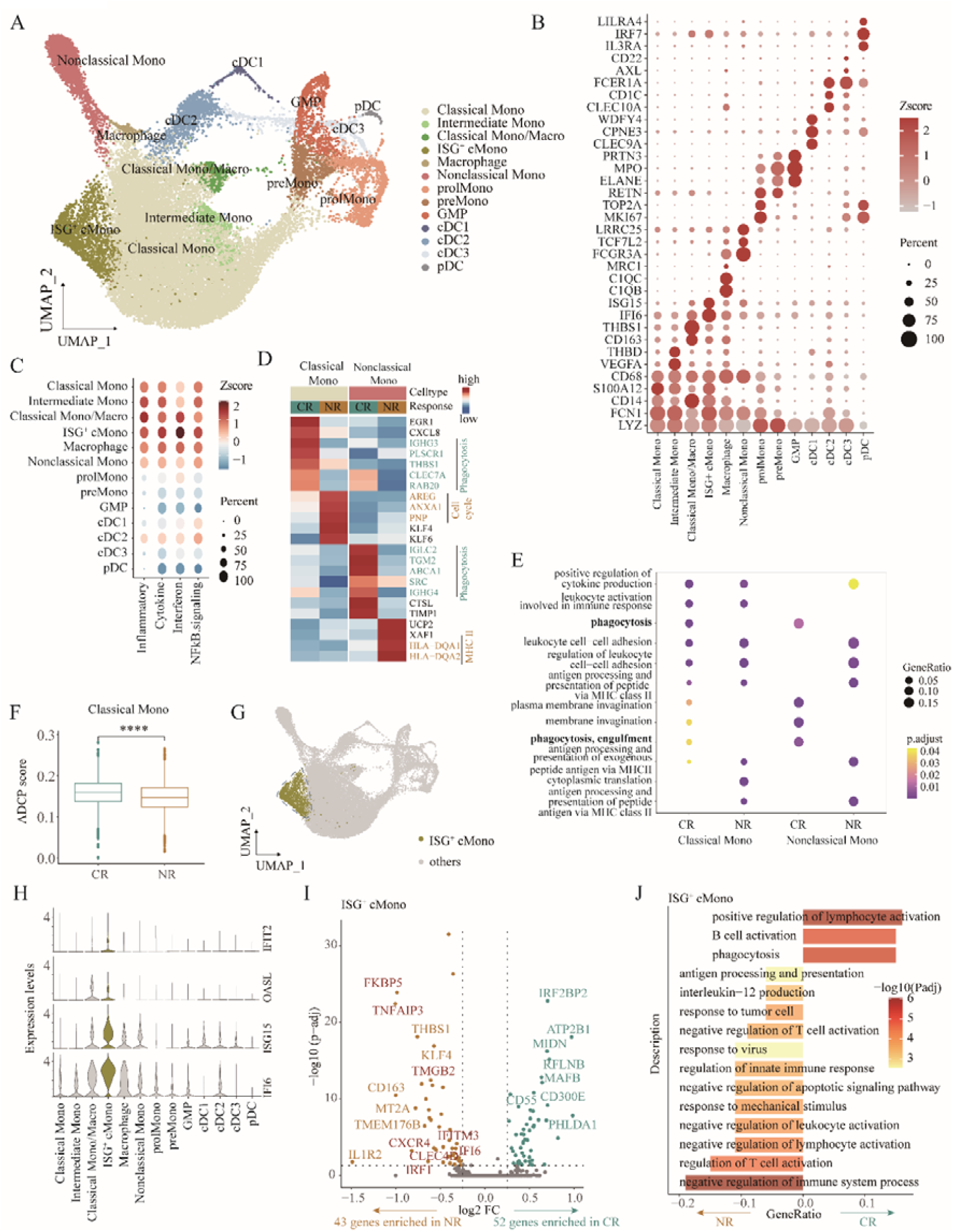
ADCP is significantly enriched in the myeloid cell clusters of the CR group. (a) Distribution of 14 myeloid cell clusters visualized on the UMAP plot. (b) Dot plot showing the expression patterns of marker genes in each myeloid cell subpopulation. (c) Dot plot displaying inflammatory, cytokine, interferon, and NF-kB signaling scores across myeloid cell subpopulations. Darker colors indicate higher scores, and larger dots represent a higher proportion of cells expressing the relevant genes within each cluster. (d) Heatmap highlighting genes with high expression in the CR and NR groups. Notably, CR-enriched genes in classical and nonclassical monocytes are associated with phagocytosis. (e) Multi-group GO enrichment analysis identifies pathways enriched in classical and nonclassical monocytes for the CR and NR groups. CR-specific pathways are mainly associated with phagocytosis. (f) Boxplot showing significant enrichment of phagocytosis-related gene sets in the CR group. (g) UMAP visualization identifies an interferon-related subset of classical monocytes, termed ISG^+^ cMono (highlighted in yellow). (h) Violin plot illustrating genes highly expressed in the ISG^+^ cMono subset. (i) High-expression genes in the ISG^+^ cMono subset are stratified between the NR and CR groups. Key enriched gene sets are labeled, with NR-enriched genes related to NF-kB signaling and inflammatory pathways highlighted in red. (j) GO enrichment analysis comparing significantly expressed genes in the ISG^+^ cMono subset between the CR and NR groups. Pathways with GeneRatio < 0 are enriched in the NR group, while those with GeneRatio > 0 are enriched in the CR group.

Using RNA velocity analysis, we found that both intermediate mono and classical mono/macro clusters were in the intermediate state of differentiation, while TAM1 were at the terminal stage of differentiation (**Supp. Fig. 2c**). *TOP2A* and *MKI67* were found in proliferating monocyte (**Fig. 2b**). Two clusters exhibited both *ELANE* features, with one cluster high in *RETN* expression defined as monocyte precursor (preMono) and the other cluster as granulocyte-monocyte progenitor (GMP). Four dendritic cell (DC) clusters were also identified, including plasmacytoid DCs (pDC) with high *IL3RA* expression, and conventional DCs of types 1, 2 and 3 (cDC1, cDC2, cDC3) expressing high levels of *ITGAX* were characterized by specific expression of *CLEC9A*, *CD1C* and *CD22*, respectively (**Fig. 2b**).

Single cell analysis revealed that classical monocytes exhibited heightened inflammatory activity, cytokine signaling and NF-kB pathway activation (**Fig. 2c**). Further analysis revealed that phagocytosis-related genes and pathway were highly enriched in both classical and nonclassical monocyte subsets in the CR group (**Fig. 2d-e**). The significantly higher phagocytosis score observed in the CR group further highlights the pivotal role of classical monocytes in facilitating daratumumab’s therapeutic efficacy through ADCP in the CR group (**Fig. 2f**).

Within the classical monocyte cluster, we identified an ISG^+^ classical Mono (cMono) subset enriched for interferon-inducible genes (e.g., *IFI6*, *ISG15*, and *IFIT2*) with notably high interferon scores (**Fig. 2c**, **Fig. 2g-h**). Differential expression analysis further confirmed that ISG^+^ cMono cells in the NR group exhibited an inflammatory phenotype, such as up-regulation of NF-kB signaling genes (*FKBP5* and *TNFAIP3*), and high expression of high inflammatory factors such as *CXCR4*, *IFITM3*, *IFI6*, and *IRF1* (**Fig. 2i**). Pathway analysis of these differentially expressed genes revealed distinct functional associations: genes related to phagocytosis and positive T cell stimulation were linked to the CR group, whereas genes involved in the negative regulation of T cell responses and the production of key immunoregulatory factors such as *IL12* were enriched in the NR group (**Fig. 2j**). These findings underscore the critical role of inflammatory myeloid cells in shaping drug responses.

### Inflammatory-associated NK cells have impaired cytotoxic capacities in NR patients

We next explored the role of NK cells in predicting the response to DR. NK cells can be functionally categorized into cytotoxic CD56^dim^ and cytokine-producing CD56^bright^ subsets. Based on their transcriptional features, we defined clusters for these subsets (**Fig. 3a**). The CD56^dim^ subset, referred to as NKdim, was characterized by elevated expression of *FCGR3A* (encoding CD16), *GZMB*, and *PRF1*, whereas the CD56^bright^ (NKbright) subset demonstrated high expression of *GZMK* and *NCAM1* (encoding CD56) (**Fig. 3b**). Differential analysis of these two NK clusters revealed that *FKBP5*, which promotes NF-kB signaling, was significantly enriched in the NR group (**Fig. 3b**). In contrast, the CR group exhibited enrichment of granzymes (*GZMA* and *GZMK*) which are known to induce cancer cell apoptosis, along with several chemokines (*CX3CR1*, *CCL3*, *CCL4* and *CCL5*) (**Fig. 3c**). Functional enrichment analysis of the NKdim group indicated that the ADCC effect was effective only in the CR patients, while the NR patients showed decreased ADCC function (**Fig. 3d-e**). Moreover, the enrichment of the cytotoxic NKbright group indicated that CR patients experienced positively mediated cytotoxicity effects, whereas cytotoxicity was suppressed in NR patients, reflecting the differential function of NK cells in the two response groups.

**Figure 3.**
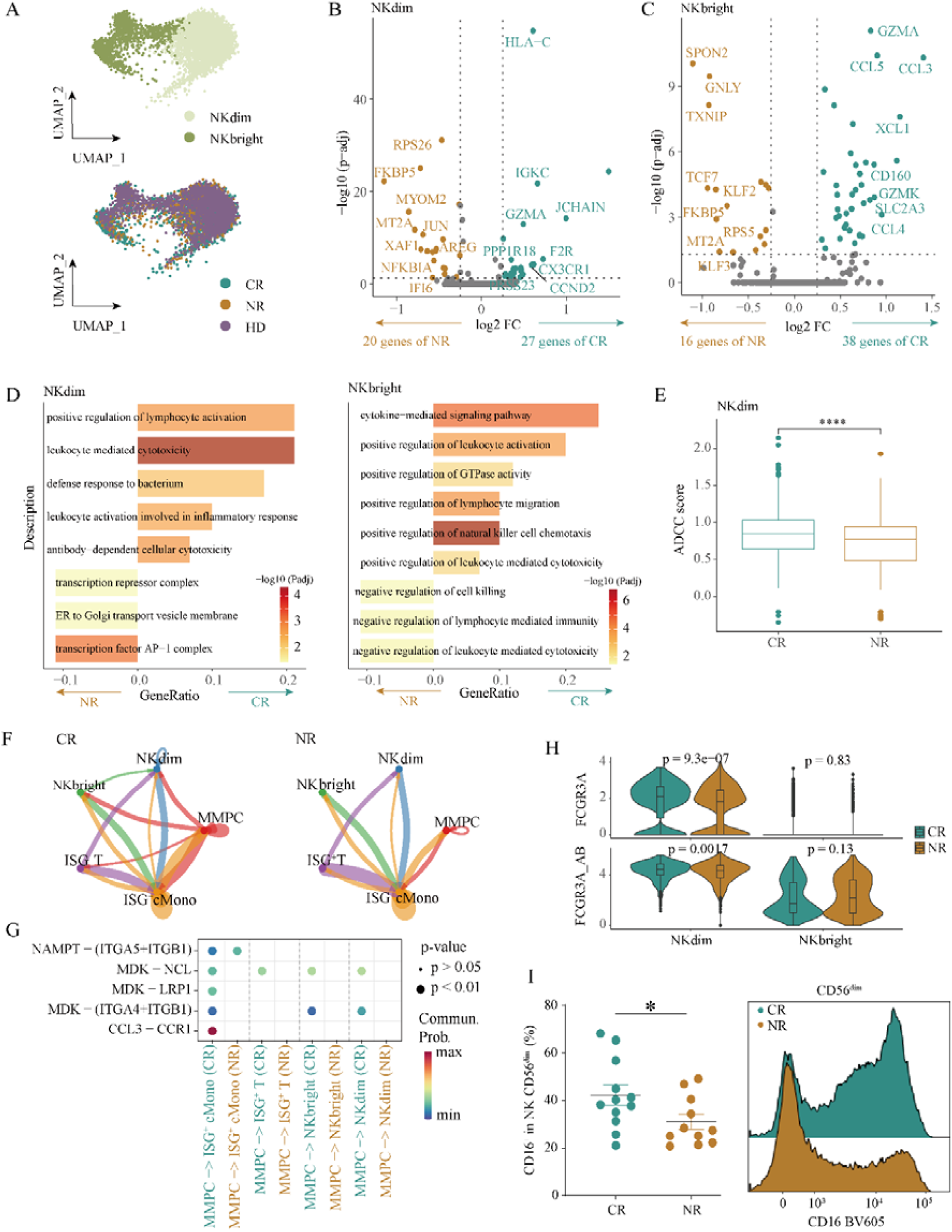
ADCC in NKdim promotes drug response. (a) UMAP plot showing the two-dimensional spatial distribution of NK cell clusters. (b-d) Differential gene expression and GO enrichment analysis of NKdim and NKbright clusters across response groups. MHC genes, chemokines (e.g., HLA-C, CCL3-5), and ADCC pathways are enriched in the CR group, whereas the NR group is associated with NF-kB genes and negative immune pathways. (e) Boxplots showing significantly higher ADCC scores in the CR group compared to the NR group within the NKdim. (f-g) CellChat analysis of ligand-receptor interactions between MMPC and immune cell populations. The CR group exhibits a higher number of interactions than the NR group, with key interactions involving CCL3-CCR1. (h) Expression levels and protein validation of the key ADCC receptor FCGR3A. (i) Percentage of CD16^+^ cells among CD56^dim^ NK cells from bone marrow aspirates. Mean fluorescence intensity of CD16 expression in CD56^dim^ NK cells. Graphs are mean ± sem. Statistical significance was determined using two-way ANOVA with Sidak’s unpaired t-test. * p<0.05.

Cellchat analysis further revealed that MMPCs in the CR patients engaged interactions with both NKdim and NKbright cells, whereas NK cells in the NR group lacked ligand-receptor interactions with MMPCs (**Fig. 3f-g**). Additionally, we observed significantly lower CD16 expression in NKdim cells at both the gene and protein levels in the NR group (**Fig. 3h**). Flow cytometry on matched samples validated this finding (**Fig. 3i and Supp. Fig. 3**). This data suggests that reduced CD16 expression on NKdim cells in NR patients may impair their ability to interact with tumor cells, thereby impairing their cytotoxic activity.

### Distinct T cell profiles between NR patients and CR patients

T cells play a crucial role in adaptive immunity and are essential for the successful elimination of tumor cells by the immune system(17). To dissect the diversity of T cells in MM, we further extracted and clustered T cells and observed five conventional CD4^+^ clusters (CD4 naïve, CD4 central memory, CD4 effector memory), three CD8^+^ clusters (CD8 naïve, CD8 memory, CD8 cytotoxic), regulatory T (Treg) cells, T helper 17 cells (Th17), ISG^+^ T cells, γδT, NKT cells, and two NK clusters (NK bright and NK dim) (**Fig. 4a-c and Supp. Fig. 4a-d**). Most effector T cell populations had higher activity (effector scores) in the CR group compared to NR (**Fig. 4d**).

**Figure 4.**
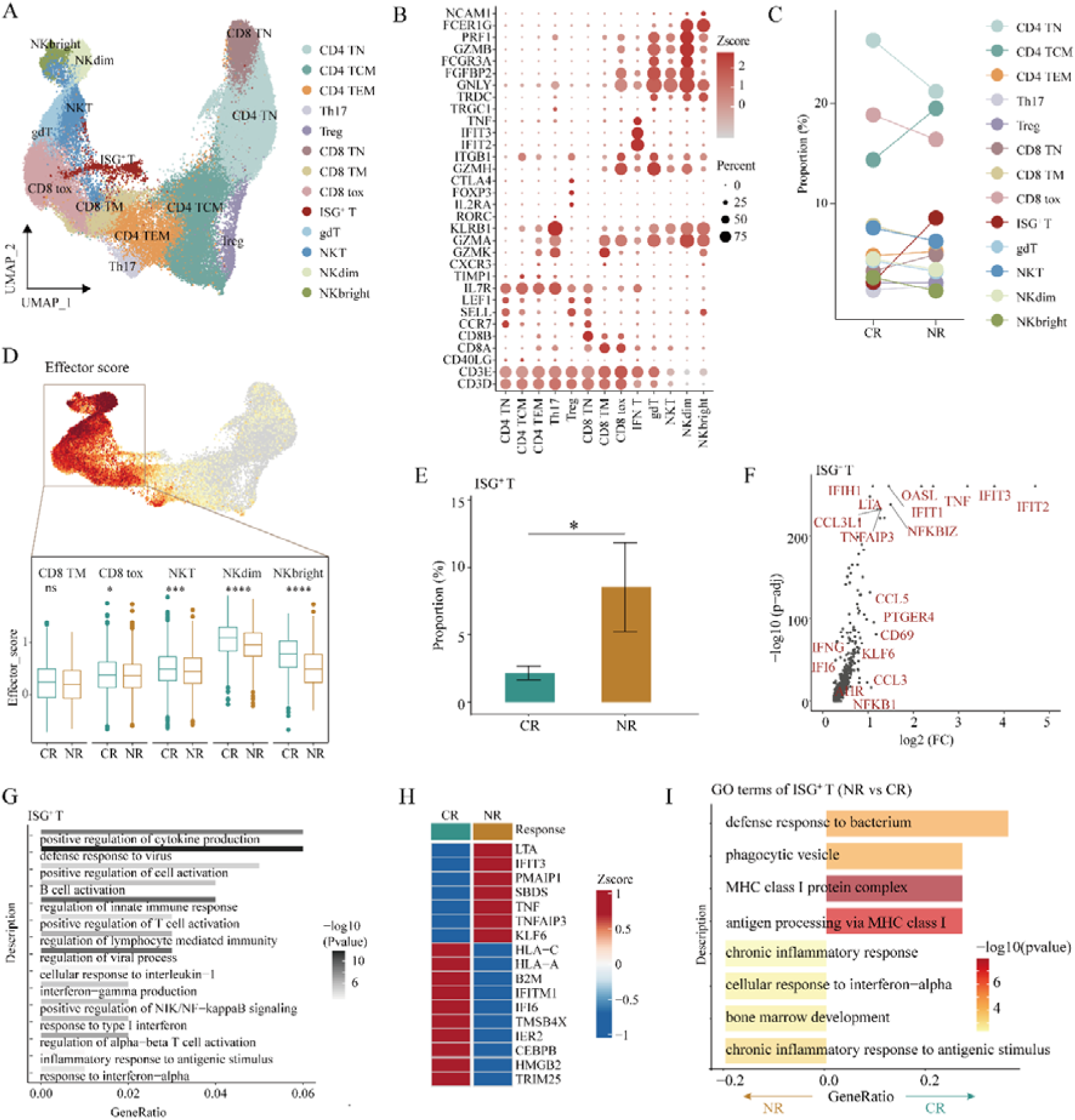
T cell subpopulation influences drug response by modulating the inflammatory status of the immune microenvironment. (a) UMAP plot showing the distribution of T cell subclusters. (b) Dot plot displaying marker expression levels across T cell subpopulations. (c) Bar graphs illustrate proportional differences in T cell subpopulations across response groups. (d) The upper panel shows feature plots of effector gene scores across T cell subpopulations, while the lower panel presents boxplots comparing effector scores between response groups for clusters with high effector activity. The CR group exhibits significantly higher effector scores, indicating a greater abundance of activated T cell populations. (e) The ISG^+^ T subpopulation is significantly enriched in the NR group. (f) Genes highly expressed in ISG^+^ T cells are highlighted, with inflammatory genes marked in red. (g) Enriched pathways associated with the highly expressed genes in ISG^+^ T cells are presented. (h) Differential gene expression analysis of ISG^+^ T cells between response groups reveals that the NR group shows elevated expression of inflammation-related genes, such as *LTA*, *IFIT3*, and *TNF*. (i) GO enrichment analysis of highly expressed genes in ISG^+^ T cells across response groups highlights distinct pathways for CR and NR groups.

We found that ISG^+^ T cells cluster, defined by expression of *TNF*, *IFIT1*, and *IFIT2*, had higher scores for inflammation, cytokine, and interferon signaling compared to most other T cell clusters (**Supp. Fig. 4e**). Interestingly, ISG^+^ T cells were more abundant in NR (**Fig. 4e**). This cell population exhibited high expression of pro-inflammatory genes, including *LTA*, *CCL3L1*, *CCL5*, *KLF6*, and *IFNG*, and were enriched in inflammatory pathways, including IL1B, IFN-γ, and NF-kB, all key regulators of immune and inflammatory responses (**Fig. 4e-g**). Additionally, the NR group showed higher expression of inflammatory factors and greater enrichment in inflammation-related pathways (**Fig. 4h-i**). These findings highlight the distinct T cell phenotypes associated with complete response and non-response to treatment in MM. The enrichment of activated CD8 memory and cytotoxic cells in CR suggests their potential role in promoting effective anti-tumor immune responses. In contrast, the higher proportion of pro-inflammatory ISG^+^ T cells in NR could contribute to a dysfunctional immune response.

### Inflammatory microenvironment correlates with NF-kB activation in MM plasma cells

The bulk RNA sequencing (RNA-seq) of tumor cells was performed in support of the single cell results. We analyzed 11 MM tumor cells (5 CR and 6 NR) by bulk RNA-seq to investigate the clinical and biological properties of CR and NR (**Fig. 5a**). Principal component analysis (PCA) showed that cancer cells from CR and NR patients were clearly separated within the first two principal components (PCs), highlighting distinct biological differences between the two groups (**Fig. 5a**). The top 100 differentially expressed genes were identified for each group, with NR highly expressing NF-kB-related genes, including *CLEC7A*, *CCR7*, *TRIM8*, *CARD9*, *SLC44A2*, *MAP3K3*, *FLOT1* and *TLR2* (**Fig. 5b**).

**Figure 5.**
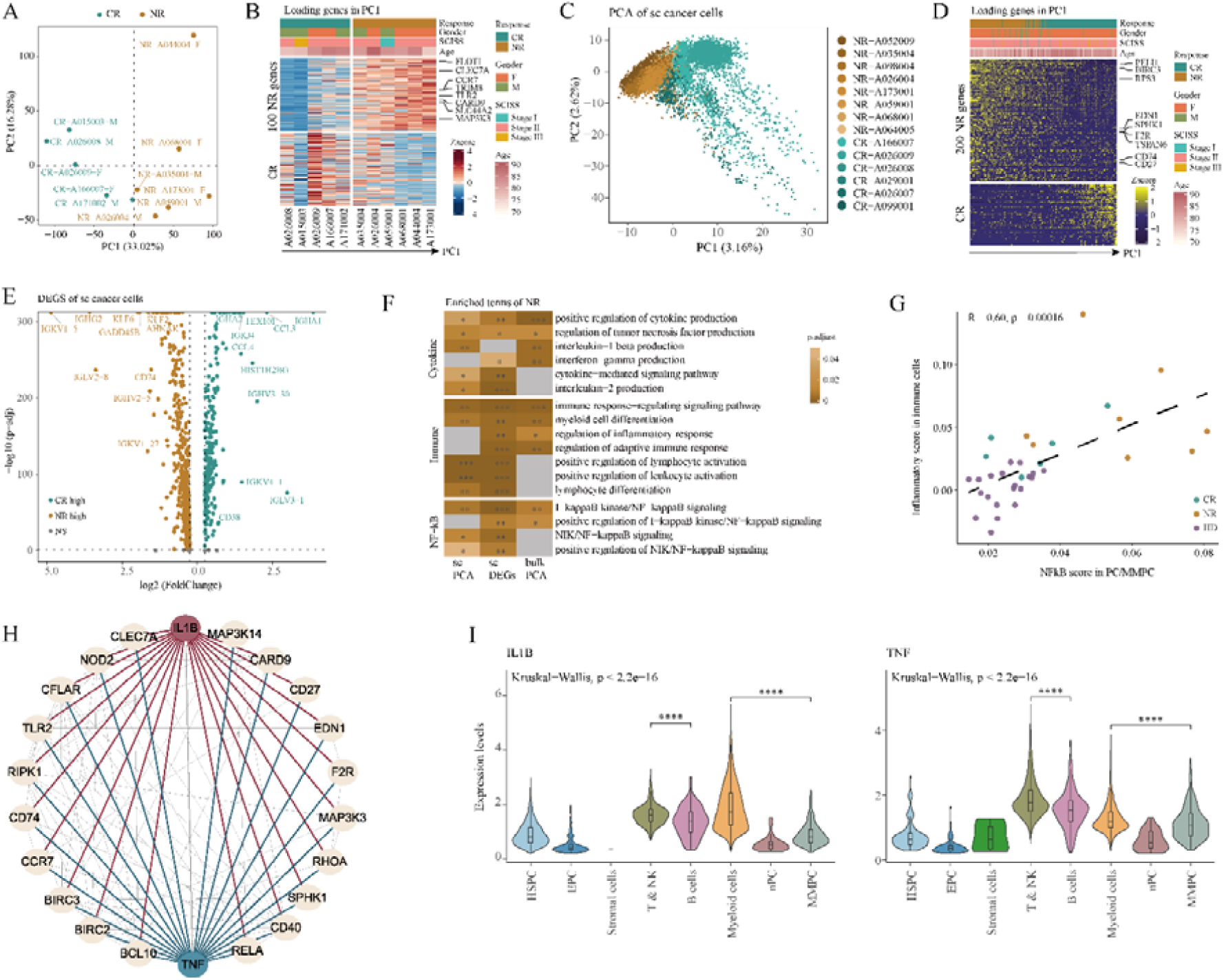
scRNA-seq and bulk RNA data analysis dissects heterogeneity of tumor cells. (a-b) PCA analysis of bulk RNA-seq data from cancer cells. (c-d) PCA analysis of scRNA-seq data from cancer cells. (e) Different expression genes analysis of scRNA-seq data from cancer cells. (f) The PCA genes from bulk/scRNA-seq data and the differential genes from scRNA-seq analysis reveal pathways that are specifically enriched in the NR group, including cytokines, immune response, and NF-kB signaling. (g) Correlation between NF-kB signatures in tumors and inflammatory signatures in the immune microenvironment. (h) Protein-protein interaction network of NF-kB - related genes regulated by key cytokines TNF and IL1B. (i) TNF and IL1B cytokines are highly expressed in immune microenvironment T and myeloid cells

We also analyzed the MMPC present in the single-cell sequencing data from 21 patients. To better characterize the tumor cells, we used 694 normal PC from HD as reference to calculating the copy number variation (CNV) scores. All MMPC had higher CNV score than the healthy referents (**Supp. Fig. 5a**), and the CNV score was significantly higher in NR patients, indicating a greater level of genomic variants in these patients (**Supp. Fig. 5a**). A total of 12,928 MMPC were identified in the single-cell sequencing data.

To explore tumor cell-specific alterations, we examined the high tumor gene scores and extracted them for re-dimensional clustering analysis. Uniform manifold approximation and projection (UMAP) showed that tumor cells were individual-specific, confirming that tumor cells in each sample were specific (**Supp. Fig. 5b-c**). Consistent with the bulk RNA-seq results, the top two PCs from scRNA-seq data clearly distinguished between cancer cells from with CR and NR patients, further illustrating distinct biological differences (**Fig. 5c**). This finding underscores the specificity between response groups, suggesting that these distinctions are more significant than individual patient variability. Importantly, when clinical factors such as gender, age, and clinical stage were considered, patient distribution along PC1 remained independent of these variables. This observation indicates that the variance observed between CR and NR patients primarily relates to drug response.

We then calculated the genes highly expressed in each response group at the PC1 level, identifying the top 200 genes in NR patients, which were associated with cytokines, immune inflammatory response and nuclear factor kappa B (NF-kB) signaling pathway (**Fig. 5d**). The NF-κB pathway regulates inflammatory genes, which influence cancer development and pathogenesis of MM (17). Further, differential gene expression analysis at the scRNA-seq level showed enrichment of important transcription factors (TF) such as *KLF6* and *KLF2* in the NR group (**Fig. 5e**). Gene ontology (GO) analysis confirmed that NR patients were characterized by immune inflammatory response, NF-kB signaling, and cytokine production (**Fig. 5f**). Pseudo-bulk analysis showed consistent results for NR (**Supp. Fig. 5d-f**).

We also identified a set of 34 genes associated with NF-kB signaling in NR patients (**Supp. Fig. 5g)**. Among these, 24 genes were linked to poor prognosis (univariate Cox P < 0.05 & hazard ratio [HR] < 1) in 853 MM, and 12 genes were also associated with poor prognosis in 172 MM treated with lenalidomide or daratumumab (**Supp. Fig. 5h-i**). The poorer prognosis in NR patients suggests that NF-kB signaling is a key driver of resistance to the combination daratumumab-lenalidomide.

We further observed a correlation between the inflammation signature of the immune microenvironment and NF-kB activation in tumor cells (**Fig. 5g**). Text mining and co-expression databases STRING revealed that cytokine *TNF* and *IL-1*β were co-regulated with NF-kB signaling genes in the tumor cells of NR patients (**Fig. 5h**). Moreover, we found *TNF* and *IL-1B* were enriched in the BME, especially in T, NK and myeloid cells (**Fig. 5i**). These findings illustrate that the enrichment of NF-kB signaling within tumor cells co-occurs with inflammatory microenvironment, contributing to drug resistance in NR patients.

### Predictive models of drug response

By performing PCA and differential analysis on bulk RNA-seq, scRNA-seq and pseudo bulk data, we identified 34 genes enriched in the NR group that positively regulate NF-kB signaling (**Supp. Fig. 5g)**. Scoring of these 34 genes at the single cell level that NR patients consistently scored higher than CR patients (**Fig. 6a**). At the all-cell level, the NR group showed significantly higher scores compared to CR (**Fig. 6b**). Survival analysis using the Multiple Myeloma Research Foundation (MMRF) CoMMpass Study data revealed that patients with high expression of these 34 genes had a significantly lower likelihood of survival (**Fig. 6c**). This finding underscores the critical role of the 34 NF-kB signaling genes in driving disease progression.

**Figure 6.**
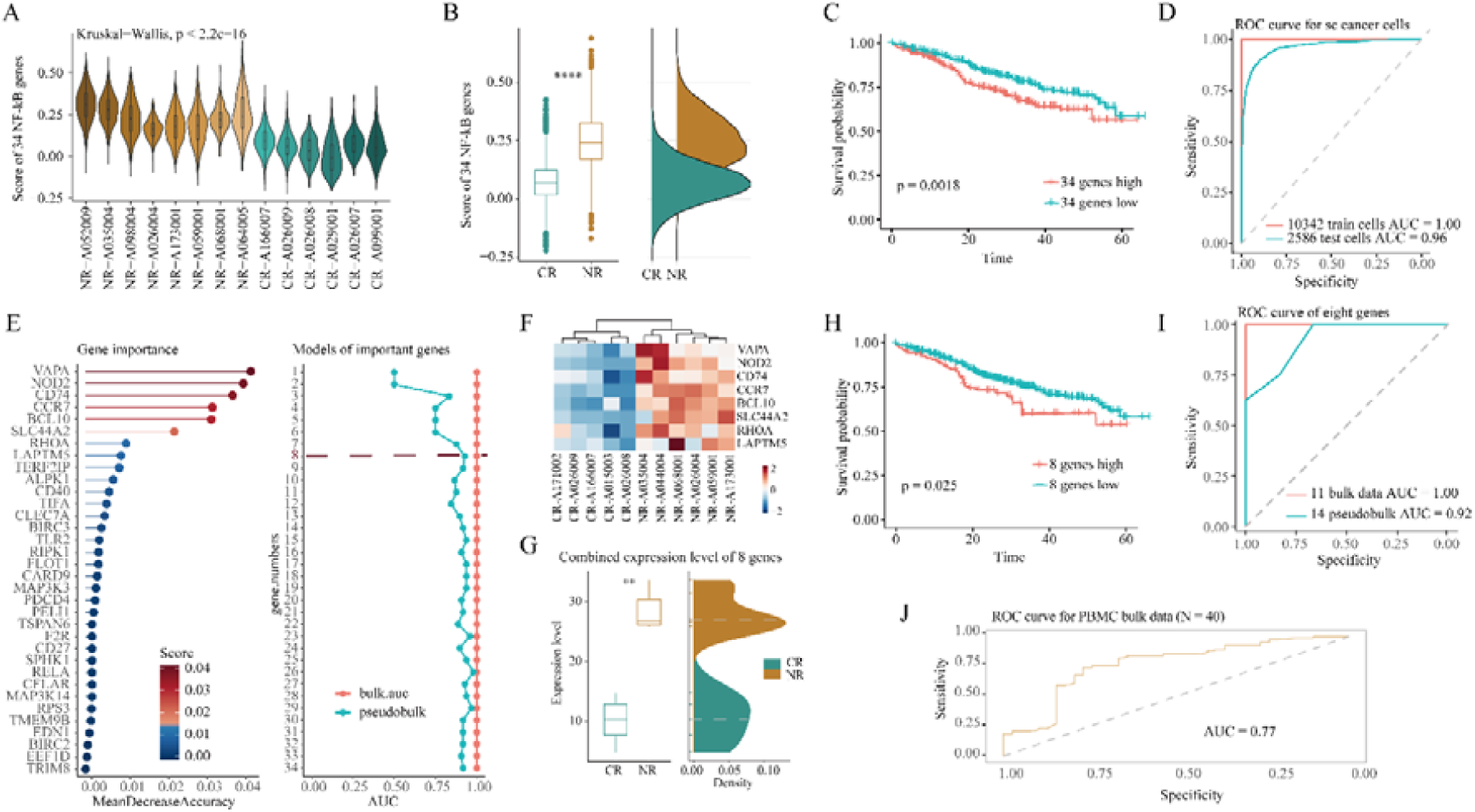
Classification model construction based on NF-kB signaling. (a-b) NF-kB genes can differentiate cell groups at the single-cell level. (c) The results of survival analysis for 34 NF-kB signaling genes using the data, where the high-expression group corresponds to a poorer prognosis. (d) The prediction results of cancer cells from single-cell sequencing using the random forest model, with an accuracy of 0.96 for 2,586 cancer cells in the test set. (e-i) The model results of the important genes identified by the random forest model in bulk-level bone marrow samples. The top 8 genes showed good predictive classification results in bulk data and pseudobulk data levels. (j) A predictive model for 40 PBMC samples was constructed based on the top 8 genes, achieving an AUC of 0.77.

To further test the predictive power of the model for the 34 genes, we conducted a random forest (RF) classifier. Using 80% tumor cells (10,342 tumor cells) as the training set and the remaining 20% cancer cells (2,586 tumor cells) as the test set, the RF classifier achieved an AUC of 0.93 on the training set. The validation set obtained an AUC value of 0.96, demonstrating high accuracy in distinguishing CR from NR patients (**Fig. 6d**).

We next ranked the candidate genes according to their importance as measured by gene importance scores at the bulk data level (**Fig. 6e**). To optimize the model, we employed a forward elimination approach, sequentially adding genes one by one, resulting in a total of 34 predictive models. Remarkably, regardless of the number of NF-kB signaling genes included, consistent high-precision was achieved, with an AUC of 1 using the leave-one-out cross-validation method on bulk data from 11 patients.

Following the principle that the optimal model should possess fewer features possible while delivering superior predictive performance, the final model was refined to include the top eight genes (*VAPA*, *NOD2*, *CD74*, *CCR7*, *BCL10, SLC44A2*, *RHOA* and *LAPTM5*), which provided the highest predictive performance while still maintaining a significant difference in survival probability (**Fig. 6f-h**). This optimized model using the eight genes demonstrated an AUC of 0.92 when predicting on a pseudo bulk test set of 14 bone marrow patients (**Fig. 6i**). Interestingly, when these 8 genes were applied to 40 peripheral blood PBMC samples from newly diagnosed MM patients, the RF model achieved robust predictive performance with an AUC value of 0.77 under 5-fold cross-validation (**Fig. 6j**). Hence, leveraging the insights from these eight screened genes, we were able to accurately predict and differentiate between CR and NR patients, providing a framework for anticipating treatment response based on NF-kB signaling.

## Discussion

In this study, we conducted a comprehensive single-cell analysis of the BME in NDMM elderly frail patients treated with a combination of DR, in the IFM2017-03 trial. We identified distinct immune and molecular differences between CR and NR. NR patients exhibiting an inflamed microenvironment characterized by elevated inflammatory cytokines and interferon signatures, predominantly within myeloid and NK cell populations. NF-kB signaling was significantly enriched in NR patients, correlating with immune dysfunction, impaired NK cell cytotoxicity, and reduced phagocytic activity of classical monocytes. Using these findings, we developed a predictive model based on NF-kB-related genes that accurately classified patients into CR or NR groups, highlighting the central role of NF-kB signaling in mediating resistance to DR therapy.

Previous studies have demonstrated the importance of the inflammatory microenvironment in the disease progression, especially from precursor stages to overt MM (18). Here we demonstrate its critical role in drug resistance and particularly regarding monoclonal antibody-based treatments. Our findings of the upregulation of NF-kB signaling in NR patients are also supported by existing literature, where this pathway has been implicated in promoting cancer cell proliferation, survival, and resistance to apoptosis. Additionally, the reduced functionality of NK cells in NR patients, particularly the decreased expression of CD16 on cytotoxic CD56^dim^ (NKdim) cells, is in line with previous reports of impaired NK cell activity in MM patients compared to healthy donors(13).

One critical question that remains unclear is whether the activation of NF-kB in MM cells precedes or follows the development of an inflammatory microenvironment. It is unclear whether NF-kB activation in tumor cells initiates the release of pro-inflammatory cytokines, such as TNF and IL-1β, which subsequently shape the immune landscape, or whether the inflamed microenvironment itself triggers NF-kB activation within the tumor cells. Some studies have suggested that tumor cells, through NF-kB activation, can induce immune cells towards a pro-tumorigenic phenotype(19,20), while others propose that an already inflamed microenvironment can drive NF-kB activation in tumor cells, promoting resistance to treatment(21,22). Future studies in precursor stages could help to resolve this question. Given the observed inflammatory microenvironment in NR patients, the well-established anti-inflammatory properties of dexamethasone—particularly its inhibition of NF-κB activation and pro-inflammatory cytokine production(23)—may function as a critical modulator of treatment efficacy. This suggests that inflammatory profiling could inform optimal corticosteroid dosing strategies within DR-containing regimens to maximize therapeutic outcomes.

Our predictive model, which incorporated key NF-kB signaling genes, achieved high accuracy in identifying NR patients, paving the way for developing tools to enable personalized treatment strategies. Beyond predicting responses, another approach could involve targeting inflammation pathways - through cytokines or NF-kB activation - to improve the efficacy of MM treatment. Blocking pro-inflammatory cytokines such as TNF or IL-1β or directly inhibiting NF-kB could be synergistic to MM treatment combination such as daratumumab and lenalidomide.

While our study primarily focused on multiple myeloma, the role of interferon signaling in driving resistance to immunotherapy has been extensively studied in various cancer types, particularly in solid tumors. Several studies have shown that chronic type I interferon signaling can promote resistance to checkpoint blockade immunotherapy (24,25) and that tumor-intrinsic type I interferon signaling can induce a T cell-low state associated with resistance to immunotherapy(26,27). Persistent STAT1 activation or sustained type I interferon signaling can inhibit NK cell proliferation and functional activities including cytolytic capacity and IFN-γ production, which could further compromise NK cells mediated anti-tumor immune responses(28). Although these studies were primarily conducted in preclinical solid tumor models, the high levels of interferon signaling observed in myeloid cells of NR patients in our study suggest that similar mechanisms may be at play in multiple myeloma, underscoring the importance of the inflamed microenvironment in driving resistance to immunotherapy across different cancer types(28,29).

Despite the valuable insights provided by our study, several limitations should be acknowledged. First, while our sample size was adequate to detect significant differences, it remains relatively small in the context of the complex and heterogeneous nature of multiple myeloma. Expanding the cohort in future studies will be essential to further validate our findings and refine the predictive model. Second, our study focused on the pretreatment bone marrow context, therefore it does fully reflect the dynamic interactions between immune and tumor cells throughout the course of treatment. To gain a more comprehensive understanding of immune responses to daratumumab and lenalidomide therapy, longitudinal sampling of the BME would be required. Finally, our analysis centered on the immune microenvironment, therefore future studies should integrate additional approaches, such as genomic or epigenetic profiling, to uncover the broader regulatory mechanisms underlying NF-kB-mediated drug resistance and identify novel therapeutic targets.

In conclusion, our study provides novel insights into the role of the inflammatory microenvironment and NF-kB signaling in mediating resistance to DR therapy in MM. By identifying key immune cell dysfunctions and developing a predictive model based on NF-kB-related genes, we have laid the foundation for the development of more personalized therapeutic strategies aimed at overcoming drug resistance in poor responders.

## Methods and materials

### Sample preparation for single-cell RNA sequencing

Immune cells were enriched from fresh BM samples by Ficoll separation and then frozen in FBS supplemented with 10% DMSO until further processing. Frozen samples were thawed at 37°C and diluted in RPMI supplemented with 20% FBS and 10 mg/ml DNAse I (StemCell). Cells were pelleted for 10 min at 300 g and resuspended in PBS/ 0.5% BSA. Dead cells were removed using Dead Cell Removal Kit (Miltenyi) following manufacturer instructions. Live cell fraction was subsequently incubated with 20 ml anti-CD138 and 20 ml anti-CD235a MicroBeads (Miltenyi). CD138+CD235+ and CD138-CD235a-fractions were obtained through magnetic isolation using MS columns (Miltenyi) following manufacturer instructions. Cell viability was checked by Calcein/Draq7 by flow cytometry and the CD138-CD235a-was then labelled with antibodies for BD Rhapsody following manufacturer’s instructions. Briefly, cells were incubated with Human BD Fc Block and then co-labelled with a Sample Tag and 35 AbSeq antibodies for surface protein expression profiling according to the manufacturer’s recommendations (Single Cell Labelling with the BD™ Single-Cell Multiplexing Kit and BD™ AbSeq Ab-Oligos Doc ID: 214419 Rev. 1.0). The AbSeq, specific for the following markers were used: CD127 (BD Biosciences Cat# 940012, RRID:AB_2875903), CD19 (BD Biosciences Cat# 940247, RRID:AB_2876128), CD45 (BD Biosciences Cat# 940002, RRID:AB_2875893), CD38 (BD Biosciences Cat# 940013, RRID:AB_2875904), CD11c (BD Biosciences Cat# 940265, RRID:AB_2876144), IgD (BD Biosciences Cat# 940026, RRID:AB_2875917), CD123 (BD Biosciences Cat# 940020, RRID:AB_2875911), CD16 (BD Biosciences Cat# 940006, RRID:AB_2875897), CD194 (BD Biosciences Cat# 940047, RRID:AB_2875938), CD3 (BD Biosciences Cat# 940000, RRID:AB_2875891), CD45RA (BD Biosciences Cat# 940011, RRID:AB_2875902), CD27 (BD Biosciences Cat# 940018, RRID:AB_2875909), CD28 (BD Biosciences Cat# 940017, RRID:AB_2875908), CD183 (BD Biosciences Cat# 940030, RRID:AB_2875921), CD161 (BD Biosciences Cat# 940283, RRID:AB_2876160), CD185 (BD Biosciences Cat# 940042, RRID:AB_2875933), CD24 (BD Biosciences Cat# 940028, RRID:AB_2875919), CD45RO (BD Biosciences Cat# 940022, RRID:AB_2875913), CD197 (BD Biosciences Cat# 940014, RRID:AB_2875905), CD8 (BD Biosciences Cat# 940003, RRID:AB_2875894), CD25 (BD Biosciences Cat# 940463, RRID:AB_2876314), CD20 (BD Biosciences Cat# 940016, RRID:AB_2875907), HLA-DR (BD Biosciences Cat# 940010, RRID:AB_2875901), CD4 (BD Biosciences Cat# 940001, RRID:AB_2875892), CD56 (BD Biosciences Cat# 940007, RRID:AB_2875898), CD196 (BD Biosciences Cat# 940033, RRID:AB_2875924), CD14 (BD Biosciences Cat# 940005, RRID:AB_2875896), CD138 (BD Biosciences Cat# 940213, RRID:AB_2876095), CD34 (BD Biosciences Cat# 940264, RRID:AB_2876143), CD279 (BD Biosciences Cat# 940015, RRID:AB_2875906), TIM-3 (BD Biosciences Cat# 940066, RRID:AB_2875957), LAG-3 (BD Biosciences Cat# 940080, RRID:AB_2875971), CD152 (BD Biosciences Cat# 940034, RRID:AB_2875925), CD11b (BD Biosciences Cat# 940008, RRID:AB_2875899), TCRgd (BD Biosciences Cat# 940057, RRID:AB_2875948). After labelling and washing steps, cells were stained with viability markers (Calcein AM - Thermo Fisher Scientific Cat. No. C1430; Draq7 - BD Biosciences Cat. No. 564904) to estimate cell concentration and viability using the BD Rhapsody™ Scanner (BD Biosciences, La Jolla, CA, USA). The viability was estimated to be between 64 and 93% (average of 83%).

### Single-cell capture and cDNA synthesis

The Single-cell isolation using the BD Rhapsody™ platform and cDNA preparation were performed according to the manufacturer’s instructions (Single Cell Capture and cDNA Synthesis with the BD Rhapsody™ Single-Cell Analysis System Doc ID: 210966 Rev. 1.0). The use of a unique sample Tag allows to combine and load multiple samples on a single BD Rhapsody cartridge. In this study, three samples were loaded per cartridge to reach the target of 8000 cells per sample, and seven cartridges were used to analyze the 21 samples selected. After quantity and quality controls, labelled samples were pooled in equal proportions in the cold Sample Buffer (BD Rhapsody Cat. No. 650000062) for a total of 35000 cells. Single cells were isolated in cartridge microwells, with magnetic beads for poly-A based mRNA capture. At loading step, the Rhapsody scanner detected, on average for the seven cartridges, 28000 wells with viable cells and 23000 wells with viable cells and a magnetic bead, which theoretically corresponds to 7700 cells per sample, a value close to our target. After cell lysis, beads were magnetically retrieved from the microwells, pooled into a single tube before reversing transcription of the poly-A captured mRNA, AbSeq and Sample Tag.

### Preparation of the whole transcriptome mRNA, AbSeq and Sample Tag libraries

After cDNA synthesis, two parallel workflows were performed following the manufacturer’s protocol (mRNA Whole Transcriptome Analysis (WTA), AbSeq, and Sample Tag Library Preparation Protocol 23-21752-00 10/2019), one for the whole transcriptome libraries using a random priming approach, and another one for the Abseq and Sample tag libraries based on PCR amplifications. Final libraries were quantified with the Qubit dsDNA HS assay kit (Invitrogen Cat. No. Q32851) and the quality was analyzed with a BioAnalyzer instrument (Agilent Technologies, RRID SCR_018043) using a High Sensitivity DNA chip (Agilent Technologies Cat. No. 5067-4626). Before pooling, libraries concentration was normalized to 4nM.

### Library pooling and sequencing

WTA, AbSeq and Sample Tag libraries were pooled and sequenced on the NovaSeq™ 6000 sequencer (Illumina, San Diego, California, United States) using the NovaSeq 6000 S4 Reagent Kit v1.5 (200 cycles) (Illumina, Cat. No. 20028313) with a loading concentration of 300pM and the following configuration: 75bp for Read 1, 75bp for Read 2 and 8bp for index. For pooling, library ratio was calculated to target a sequencing depth of 50000 reads/cell for WTA, 17500 reads/cell for AbSeq and 600 reads/cell for Sample tag.

### Sample preparation for bulk RNAseq

CD138+ cells were isolated from BM aspirates using anti-CD138 MicroBeads (Miltenyi), then RNA was extracted. Library preparation and sequencing were performed by Novogene.

### Flow cytometry analysis of BM aspirates

Samples (12 CR and 11 NR) were thawed at 37°C and 2×10^6^ cells were stained with 1:1,000 with Zombie Aqua Fixable Viability Kit (BioLegend, cat. 423101) for 20 min at 4 °C. Fc receptors were blocked using human TruStain FcX (BioLegend, cat. 422301) following manufacturer’s instructions. Samples were then fixed with 0.2% formaldehyde for 20 min at 4 °C and stained overnight in Permeabilization buffer (eBiosciences, cat. 00-8333-56) as described by Whyte CE et al.,(29).

The following antibodies were used for staining at 1:500 final concentration: anti-human CD45 APC-Cy7 (clone 2D1, BioLegend, cat. 368515, RRID AB_2566375), anti-human CD3 BUV395 (clone UCHT1, BD Biosciences, cat. 563546, RRID AB_2744387), anti-human CD56 FITC (clone HCD56; BioLegend; cat. 318304, RRID AB_604100), and anti-human CD16 BV 605 (clone 3G8, BioLegend; cat. 302039, RRID AB_2561354). Samples were acquired using LSRFortessa X-20 (BD Biosciences) and analysis was performed using OMIQ (Dotmatics).

### Bulk RNA-seq data analyses

The bulk RNA-seq data sequencing counts of each sample were greater than 20 million. Raw counts were utilized for normalization, PCA and enrichment analysis. PCA analysis was based on a normalized matrix of counts per million (CPM) displaying a two-dimensional plane of PC1 and PC2 coordinates. Heat maps were presented using the Complex Heatmap package (version 2.12.1) with specific loading genes in order of PC1 coordinates for each patient, including 100 responder group genes and 100 non-responder group genes, respectively. The specific genes were then enriched separately by the cluster Profiler package (version 4.4.4), and the highly enriched entries that were presented met a significant level of q value less than 0.05 including at least four genes.

### Single-cell RNA sequencing analyses

Raw fastq data were mapped to the human reference genome (GRCh38) via the standard BD Rhapsody workflow according to the manufacturer’s recommendations. Unique molecular identifier (UMI) count matrices were imported into Seurat (R package, version 4.2.1 and version 4.1.1) (30)for quality control that included selecting cells with a library complexity of more than 500 features, less than 50,000 counts, filtering out cells with high percentages of mitochondrial genes (> 25%) and removing multiple and undetermined cells. To account for differences in sequencing depth across cells, RNA UMI counts were log-normalized, while antibody UMI counts were applied to a centered log ratio transformation to account for unspecific binding background signal.

### Healthy BM data

To provide additional comparisons for the patients collected in this study including the responder and non-responder groups, scRNA-seq data including 20 healthy individuals published by Oetjen et al. were collected and analyzed, and the original sequencing files were downloaded from GEO (GSE120221)(15).

### scRNA-seq data integration

scRNA-seq dataset of 21 patients and 20 normal bone marrow samples were merged by integration, normalized and analyzed by PCA across all cells using the Seurat (v4.1.1, RRID SCR_016341) R package (30). We controlled batch effects using FindIntegrationAnchors function for identify anchors and IntegrateData function for integration dataset base on anchors, the top 2000 variable genes from each dataset and the first 30 PCs were used for integration. Based on the integration results, the top 20 PCs were clustered unsupervised, using a shared nearest neighbor modular optimized clustering algorithm integrated in Seurat (resolutions 1-2), and cells were projected in two-dimensional coordinates of UMAP.

### Cell type annotation

Annotation of cell types was performed based on manual review of marker genes (as above). The subgroup clustering results were decisive for the identification of cell type identity, and mixed cell populations were focused on, on the one hand, two subgroups in a cluster due to clustering problems needed to be further subdivided and defined, and on the other hand, cell populations expressing mixed marker genes (e.g., both erythroid and immune cell features were highly expressed) were considered as mixed cells and were excluded from subsequent analysis. Cluster-specific highly expressed gene expression analyses were all performed using the FindAllMarkers function of the Seurat package. Only genes detected in the smallest fraction of 25% of cells in either of the two populations were tested. Adjusted P values < 0.05 were considered significant using the default Wilcoxon test.

### Module calculation based on cell proportions

First, the proportions of different cell subpopulations were calculated across MM patient response groups and the HD group. The table function was used to determine the proportion of each fine subpopulation within each Response group, followed by normalization. A heatmap was then generated using pheatmap to visualize the distribution of cell subpopulations under different response conditions.

Additionally, hierarchical clustering was performed on both samples and cell subpopulations, and the column order of each module was extracted based on the clustering results. Notably, Module4 was found to be significantly enriched in the NR group compared to the CR group, prompting further differential gene expression (DEG) analysis.

For DEG analysis of Module4, FindMarkers was applied to identify genes that were differentially expressed between the CR and NR groups, using the thresholds min.pct = 0.25 and logFC.threshold = 0.25. Genes with significant differential expressions (p_val_adj < 0.05 and |avg_log2FC| ≥ 0.25) were further selected. ggplot2 was used to visualize gene expression changes, highlighting genes with higher expression in either the CR or NR groups.

### GSEA-based functional enrichment analysis of Module4

Following the identification of Module4-associated differentially expressed genes, gene set enrichment analysis (GSEA) was performed to assess functional enrichment. First, genes were ranked in descending order based on avg_log2FC, and bitr was used to convert gene symbols (SYMBOL) to Entrez IDs to meet the input requirements for GSEA. Next, the Hallmark gene sets were retrieved using msigdbr (RRID SCR_022870), and GSEA was conducted using GSEA function with a significance threshold of pvalueCutoff = 0.05 to identify significantly enriched gene sets. Finally, gseaplot2 was employed to visualize the enrichment curves of the most significantly enriched Hallmark pathways, providing insights into the inflammation-related characteristics of Module4 in the NR group.

### Gene set enrichment scores

To assess the corresponding properties of cell clusters or different response groups, NF-kB signaling, cytokine and inflammation scores were evaluated for each cell with the ‘AddModuleScore’ function in Seurat. Inflammatory signature genes were obtained from gene set ‘HALLMARK_INFLAMMATORY_RESPONSE’ downloaded from Molecular Signatures Database (MSigDB, RRID_016863) (31), cytokine genes were obtained from the GO database called ‘positive regulation of cytokine production’ (GO: 0001819), and the NF-kB signaling gene set included ‘I kappaB kinase/NF-kappaB signaling’ (GO: 0007249) and ‘positive regulation of I-kappaB kinase/NF-kappaB signaling’ (GO: 0043123). Additionally, the effector gene set includes *PRF1*, *IFNG*, *NKG7*, *GZMB*, *GZMA*, *GZMK*, *GZMD*, *KLRK1*, *KLRB1*, *KLRD1*, *CTSW*, and *CST7*.

### Gene ontology and gene set enrichment analysis

The clusterProfiler package (RRID SCR_016884) was used to assess the enrichment of the top linked genomes in the gene ontology biological processes categories. Enrichment-adjusted significant categories with p-values < 0.05 were identified. CR and NR group specific genes in single cell clusters were calculated according to the FindMarkers function with a threshold of less express in 25% of cells, log2 fold change > 0.25 and adjusted P values < 0.05.

### RNA velocity and pseudo-time analysis

RNA velocity analysis was performed with scvelo (version 0.1.25, RRID SCR_018168) (32) in python. Bam files obtained from BD flow-based analysis were used to generate loom files. Briefly, after gene selection and normalization, first- and second-order moments were calculated using the ‘scv.pp.ments’ function. The complete splicing dynamics is recovered with the ‘scv.tl.recover_dynamics’ function, and the velocity values in dynamic mode are obtained with the ‘scv.tl.velocity’ function. The velocity is projected onto the diffusion map with the ‘scv.pl.velocity_embedding_stream’ function and displayed as a streamline. Visualize the spliced and unspliced phase maps of individual genes with the ‘scv.pl.velocity’ function. The pseudotime of the cell is obtained with the ‘scv.tl.recover_latent_time’ function.

### Single-cell RNA-seq correlation analysis

For each cell type of defined myeloid cells, we calculated the average expression values of all genes for each subpopulation using the AverageExpression function, and the top 2000 genes with the largest standard deviation were prioritized to calculate the Pearson correlation coefficients.

### Detection of CNVs in cancer cells

We inferred CNVs of the 21 patients by InferCNV using scRNA-seq data (33). Genes with a mean UMI higher than 0.1 were selected for calculation using plasma cells from normal samples as a reference and all plasma cells from patients as a test set, as described in InferCNV (RRID SCR_021140). In brief, the heatmap shows the variation score for each chromosome based on genomic position according to regions with a window of every 101 genes. Plasma cells from patients all exhibited large CNVs and clarified the significantly high variant status of NRs.

### Pseudobulk data analyses

Pseudobulk analysis was performed on a patient-by-patient basis by aggregating gene expression counts across all malignant plasma cells from each patient to construct a new expression matrix. First, MMPCs were extracted based on cell type annotations. To ensure robust analysis, patients with fewer than 10 detected MMPCs were excluded. For each retained patient, raw gene counts were summed across all MMPCs within that patient to generate a patient-level expression profile. To filter lowly expressed genes, only genes with CPM values greater than 1 in at least two patients were retained. The resulting count matrix was then log-transformed using log2 (CPM + 1) normalization. Subsequent PCA and enrichment analyses are consistent with the bulk data analysis process described above.

### Protein-Protein Interaction network analysis

To investigate the interaction between NF-kB-related genes and key inflammatory cytokines, *IL1B* and *TNF*, a protein-protein interaction (PPI) network was constructed. The STRING database (https://string-db.org/, RRID SCR_005223) was utilized to retrieve known and predicted protein-protein interactions. The interaction network was generated by inputting the 34 NF-kB-related genes, which were identified through a series of computational analyses, along with *IL1B* and *TNF*. A confidence score threshold of 0.4 was applied to filter for high-confidence interactions. The resulting interaction network was then imported into Cytoscape (RRID SCR_003032) for visualization and further analysis.

### Single-cell Random Forest model construction (bone marrow)

We used scRNA-seq data derived from bone marrow plasma cells of MM patients to construct a classification model based on 34 NF-kB-related genes. Data preprocessing included gene length normalization and TPM transformation. The dataset was split into a training set (80%) and a test set (20%), with response labels defined as CR and NR. A RF classifier was trained on the training set, and its performance was evaluated using ROC curves and AUC. The model demonstrated high predictive power, achieving an AUC of 1.0 on the training set and 0.96 on the test set, indicating strong generalizability.

### Bulk RNA-seq Random Forest model construction (bone marrow)

Like the single-cell analysis, we applied gene length normalization and TPM transformation before performing feature selection using the RF algorithm. Eight key genes were identified as the most informative for model construction. A predictive model was developed using bulk RNA-seq data and validated through leave-one-out cross-validation, achieving an AUC of 1.0. To further assess its robustness, the model was tested on pseudobulk samples, yielding an AUC of 0.92, demonstrating strong predictive performance.

### PBMC-based model construction for clinical applicability

To enhance the clinical applicability of our findings, we extended the model to PBMC-derived patient samples. We selected the same eight key genes identified in the BM bulk RNA-seq analysis. Using data from 40 PBMC samples, we trained a RF model and evaluated its performance through five-fold cross-validation. The model achieved an AUC of 0.77, suggesting its potential utility in peripheral blood-based patient stratification.

Across all model analyses, a consistent data preprocessing workflow was employed, including gene length normalization, TPM transformation, and RF based feature selection, ensuring methodological coherence across BM scRNA-seq data, PBMC scRNA-seq data and BM bulk RNA-seq data.

### Survival analysis

Survival data were downloaded from MMRF CoMMpass Study (RRID SCR_003721) and included a total of 853 patients with multiple myeloma and 172 patients treated with Lenalidomide or Daratumumab. R package survivor and survminer (RRID SCR_026512) were used for prognostic assessment, specifically, the obtained NF-kB signaling gene and patient survival information were used as inputs to generate patient survival curves and statistical tests for overall survival. Adjusted P values < 0.05 were considered statistically significant.

## Data availability

All single-cell RNA, surface protein, and bulk sequencing data generated in this study have been deposited in the European Genome-phenome Archive (EGA) under the reference XXXX. The publicly available scRNA-seq dataset from 20 healthy individuals (GSE120221) was used for comparison. All R code used for the analysis is available on GitHub at https://github.com/Chengwenwen1/MM-France/tree/main.

## Acknowledgements

We thank the facilities UMS2014-US41-PLBS BioImaging Center Lille, Flow Cytometry Core Facility, and Génomique Fonctionelle at Structurale (GFS), F-59000 Lille, France, for technical support. Figures 1A has been created with BioRender.com. This study was financed by Johnson and Johnson.

## Competing interests

The other authors declare no competing interests.

## Notes

### Competing Interest Statement

The authors have declared no competing interest.

### Clinical Trial

NCT03993912

### Funding Statement

This study was founded by Johnson and Johnson.

### Author Declarations

The French National Agency for the Safety of Medicines and Health (ANSM) and the Ethical Committee "Sud-Mediterranee II" (reference 219 A10) approved the protocol (ID# 19.02.28.43641). The IFM2017-03 trial (NCT03993912) and the correlative studies were conducted in accordance with the principles of the Declaration of Helsinki and the International Conference on Harmonization Good Clinical Practice guidelines. All the patients provided written informed consent. The trial was promoted by Lille University Hospital (CHU Lille) in collaboration with the IFM and with the support of Johnson and Johnson.

### Summary of Updates

Author affiliations and abstract have been updated.

## References

1. Dimopoulos MA, Oriol A, Nahi H, San-Miguel J, Bahlis NJ, Usmani SZ, et al. Daratumumab, Lenalidomide, and Dexamethasone for Multiple Myeloma. N Engl J Med 2016;375(14):1319–31 doi 10.1056/NEJMoa1607751.

2. Facon T, Kumar S, Plesner T, Orlowski RZ, Moreau P, Bahlis N, et al. Daratumumab plus Lenalidomide and Dexamethasone for Untreated Myeloma. N Engl J Med 2019;380(22):2104–15 doi 10.1056/NEJMoa1817249.

3. Facon T, Leleu X, Manier S. How I treat multiple myeloma in geriatric patients. Blood 2024;143(3):224–32 doi 10.1182/blood.2022017635.

4. Sanchez L, Wang Y, Siegel DS, Wang ML. Daratumumab: a first-in-class CD38 monoclonal antibody for the treatment of multiple myeloma. J Hematol Oncol 2016;9(1):51 doi 10.1186/s13045-016-0283-0.

5. Liu J, Xing L, Li J, Wen K, Liu N, Liu Y, et al. Epigenetic regulation of CD38/CD48 by KDM6A mediates NK cell response in multiple myeloma. Nat Commun 2024;15(1):1367 doi 10.1038/s41467-024-45561-z.

6. Krejcik J, Casneuf T, Nijhof IS, Verbist B, Bald J, Plesner T, et al. Daratumumab depletes CD38+ immune regulatory cells, promotes T-cell expansion, and skews T-cell repertoire in multiple myeloma. Blood 2016;128(3):384–94 doi 10.1182/blood-2015-12-687749.

7. Fink EC, Ebert BL. The novel mechanism of lenalidomide activity. Blood 2015;126(21):2366–9 doi 10.1182/blood-2015-07-567958.

8. Kronke J, Udeshi ND, Narla A, Grauman P, Hurst SN, McConkey M, et al. Lenalidomide causes selective degradation of IKZF1 and IKZF3 in multiple myeloma cells. Science 2014;343(6168):301-5 doi 10.1126/science.1244851.

9. Casneuf T, Adams HC, 3rd, van de Donk N, Abraham Y, Bald J, Vanhoof G, et al. Deep immune profiling of patients treated with lenalidomide and dexamethasone with or without daratumumab. Leukemia 2021;35(2):573–84 doi 10.1038/s41375-020-0855-4.

10. Schinke C, Poos AM, Bauer M, John L, Johnson S, Deshpande S, et al. Characterizing the role of the immune microenvironment in multiple myeloma progression at a single-cell level. Blood Adv 2022;6(22):5873–83 doi 10.1182/bloodadvances.2022007217.

11. Cheng Y, Sun F, Alapat DV, Wanchai V, Mery D, Siegel ER, et al. Multi-omics reveal immune microenvironment alterations in multiple myeloma and its precursor stages. Blood Cancer J 2024;14(1):194 doi 10.1038/s41408-024-01172-x.

12. Zavidij O, Haradhvala NJ, Mouhieddine TH, Sklavenitis-Pistofidis R, Cai S, Reidy M, et al. Single-cell RNA sequencing reveals compromised immune microenvironment in precursor stages of multiple myeloma. Nat Cancer 2020;1(5):493–506 doi 10.1038/s43018-020-0053-3.

13. Blanquart E, Ekren R, Rigaud B, Joubert MV, Baylot V, Daunes H, et al. NK cells with adhesion defects and reduced cytotoxic functions are associated with a poor prognosis in multiple myeloma. Blood 2024;144(12):1271–83 doi 10.1182/blood.2023023529.

14. de Jong MME, Kellermayer Z, Papazian N, Tahri S, Hofste Op Bruinink D, Hoogenboezem R, et al. The multiple myeloma microenvironment is defined by an inflammatory stromal cell landscape. Nat Immunol 2021;22(6):769–80 doi 10.1038/s41590-021-00931-3.

15. Oetjen KA, Lindblad KE, Goswami M, Gui G, Dagur PK, Lai C, et al. Human bone marrow assessment by single-cell RNA sequencing, mass cytometry, and flow cytometry. JCI Insight 2018;3(23) doi 10.1172/jci.insight.124928.

16. Tirier SM, Mallm JP, Steiger S, Poos AM, Awwad MHS, Giesen N, et al. Subclone-specific microenvironmental impact and drug response in refractory multiple myeloma revealed by single-cell transcriptomics. Nat Commun 2021;12(1):6960 doi 10.1038/s41467-021-26951-z.

17. Vrabel D, Pour L, Sevcikova S. The impact of NF-kappaB signaling on pathogenesis and current treatment strategies in multiple myeloma. Blood Rev 2019;34:56–66 doi 10.1016/j.blre.2018.11.003.

18. Zavidij O, Haradhvala NJ, Mouhieddine TH, Sklavenitis-Pistofidis R, Cai S, Reidy M, et al. Single-cell RNA sequencing reveals compromised immune microenvironment in precursor stages of multiple myeloma. Nature Cancer 2020;1(5):493–506 doi 10.1038/s43018-020-0053-3.

19. Grivennikov SI, Greten FR, Karin M. Immunity, inflammation, and cancer. Cell 2010;140(6):883–99 doi 10.1016/j.cell.2010.01.025.

20. Saccani A, Schioppa T, Porta C, Biswas SK, Nebuloni M, Vago L, et al. p50 nuclear factor-kappaB overexpression in tumor-associated macrophages inhibits M1 inflammatory responses and antitumor resistance. Cancer Res 2006;66(23):11432–40 doi 10.1158/0008-5472.CAN-06-1867.

21. Luo JL, Maeda S, Hsu LC, Yagita H, Karin M. Inhibition of NF-kappaB in cancer cells converts inflammation-induced tumor growth mediated by TNFalpha to TRAIL-mediated tumor regression. Cancer Cell 2004;6(3):297–305 doi 10.1016/j.ccr.2004.08.012.

22. Hagemann T, Lawrence T, McNeish I, Charles KA, Kulbe H, Thompson RG, et al. “Re-educating” tumor-associated macrophages by targeting NF-kappaB. J Exp Med 2008;205(6):1261–8 doi 10.1084/jem.20080108.

23. Auphan N, DiDonato JA, Rosette C, Helmberg A, Karin M. Immunosuppression by glucocorticoids: inhibition of NF-kappa B activity through induction of I kappa B synthesis. Science 1995;270(5234):286–90 doi 10.1126/science.270.5234.286.

24. Jacquelot N, Yamazaki T, Roberti MP, Duong CPM, Andrews MC, Verlingue L, et al. Sustained Type I interferon signaling as a mechanism of resistance to PD-1 blockade. Cell Res 2019;29(10):846–61 doi 10.1038/s41422-019-0224-x.

25. Zak J, Pratumchai I, Marro BS, Marquardt KL, Zavareh RB, Lairson LL, et al. JAK inhibition enhances checkpoint blockade immunotherapy in patients with Hodgkin lymphoma. Science 2024;384(6702):eade8520 doi 10.1126/science.ade8520.

26. Mathew D, Marmarelis ME, Foley C, Bauml JM, Ye D, Ghinnagow R, et al. Combined JAK inhibition and PD-1 immunotherapy for non-small cell lung cancer patients. Science 2024;384(6702):eadf1329 doi 10.1126/science.adf1329.

27. Qiu J, Xu B, Ye D, Ren D, Wang S, Benci JL, et al. Cancer cells resistant to immune checkpoint blockade acquire interferon-associated epigenetic memory to sustain T cell dysfunction. Nat Cancer 2023;4(1):43–61 doi 10.1038/s43018-022-00490-y.

28. Tabellini G, Vairo D, Scomodon O, Tamassia N, Ferraro RM, Patrizi O, et al. Impaired natural killer cell functions in patients with signal transducer and activator of transcription 1 (STAT1) gain-of-function mutations. J Allergy Clin Immunol 2017;140(2):553–64 e4 doi 10.1016/j.jaci.2016.10.051.

29. Whyte CE, Tumes DJ, Liston A, Burton OT. Do more with Less: Improving High Parameter Cytometry Through Overnight Staining. Curr Protoc 2022;2(11):e589 doi 10.1002/cpz1.589.

30. Argelaguet R, Clark SJ, Mohammed H, Stapel LC, Krueger C, Kapourani CA, et al. Multi-omics profiling of mouse gastrulation at single-cell resolution. Nature 2019;576(7787):487–91 doi 10.1038/s41586-019-1825-8.

31. Liberzon A, Subramanian A, Pinchback R, Thorvaldsdottir H, Tamayo P, Mesirov JP. Molecular signatures database (MSigDB) 3.0. Bioinformatics 2011;27(12):1739–40 doi 10.1093/bioinformatics/btr260.

32. Bergen V, Lange M, Peidli S, Wolf FA, Theis FJ. Generalizing RNA velocity to transient cell states through dynamical modeling. Nat Biotechnol 2020;38(12):1408–14 doi 10.1038/s41587-020-0591-3.

33. Tirosh I, Izar B, Prakadan SM, Wadsworth MH, 2nd, Treacy D, Trombetta JJ, et al. Dissecting the multicellular ecosystem of metastatic melanoma by single-cell RNA-seq. Science 2016;352(6282):189–96 doi 10.1126/science.aad0501.

