## Supplementary figures and legends for "Inflammatory microenvironment impairs the therapeutic effect of daratumumab-lenalidomide in multiple myeloma"

Supplementary Table 1 The main clinical characteristics of patients

| | CR (n=11) | NR (n=10) |
|---|---|---|
| Mean age | 79 (71-86) | 81 (73-89) |
| Median age | 78 | 82 |
| St. dev age | 5.6 | 4.6 |
| Females | 55% | 70% |
| Males | 45% | 30% |
| Mean sc tumor (n° samples; min – max) | 742 (8; 203-3128) | 1165 (6; 206-1921) |
| Mean sc n-tumor (min – max) | 3217 (2101-5202) | 3564 (1407-4980) |

Supplementary Table 2 The main characteristic of 20 healthy doners

| | normal BM (n=20) |
|---|---|
| Mean age | 51 (24-84) |
| Median age | 53 |
| St. dev age | 515 |
| Females | 50% |
| Males | 50% |

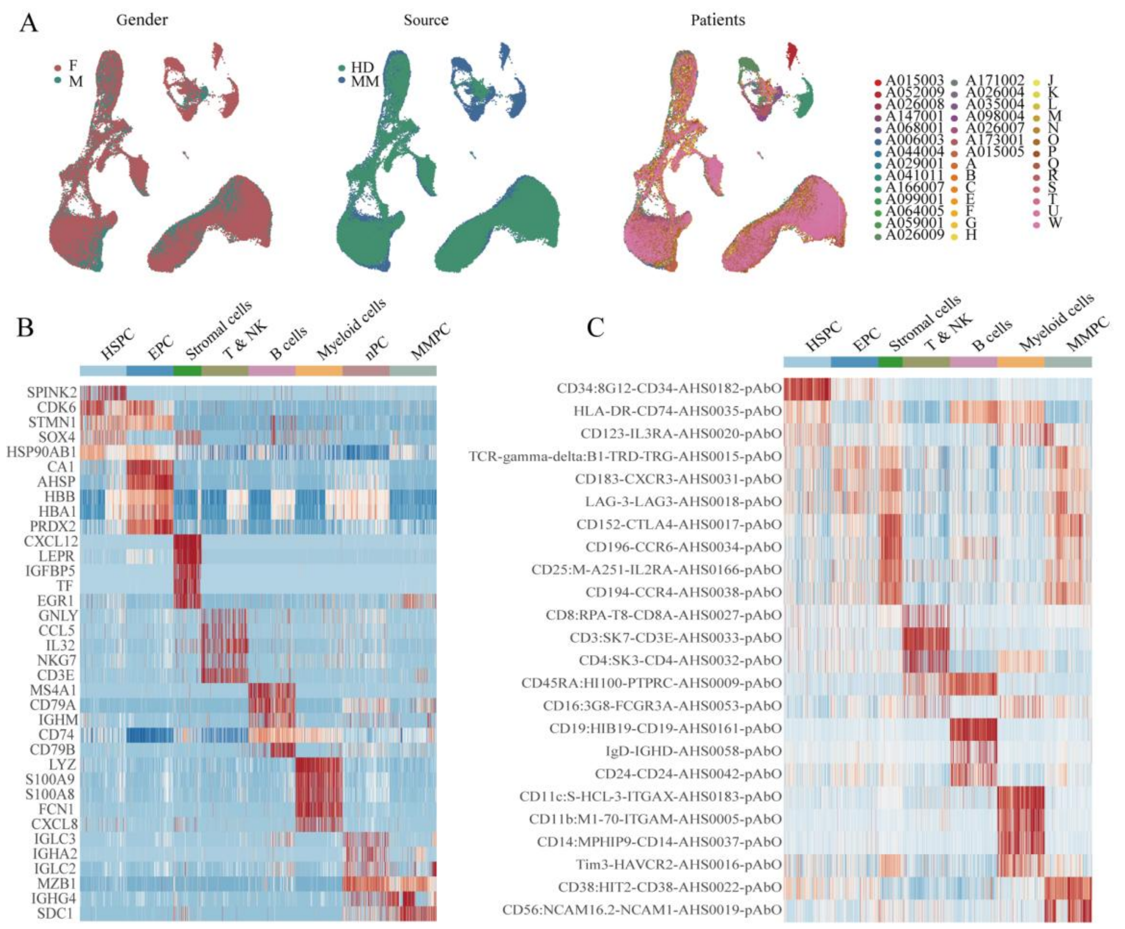


**Supplementary figure 1** Summary of the human bone marrow microenvironment atlas. (a) UMAP plots demonstrate the removal of batch effects across gender (left), health status (middle), and individual samples (right), ensuring the robustness and reliability of the atlas. (b) Heatmap displaying the top 5 highly expressed genes for each subpopulation in multiple myeloma patients. (c) Heatmap illustrates the top 3 enriched surface proteins for each subpopulation in multiple myeloma patients, based on differential analysis of 35 surface proteins measured in this study.

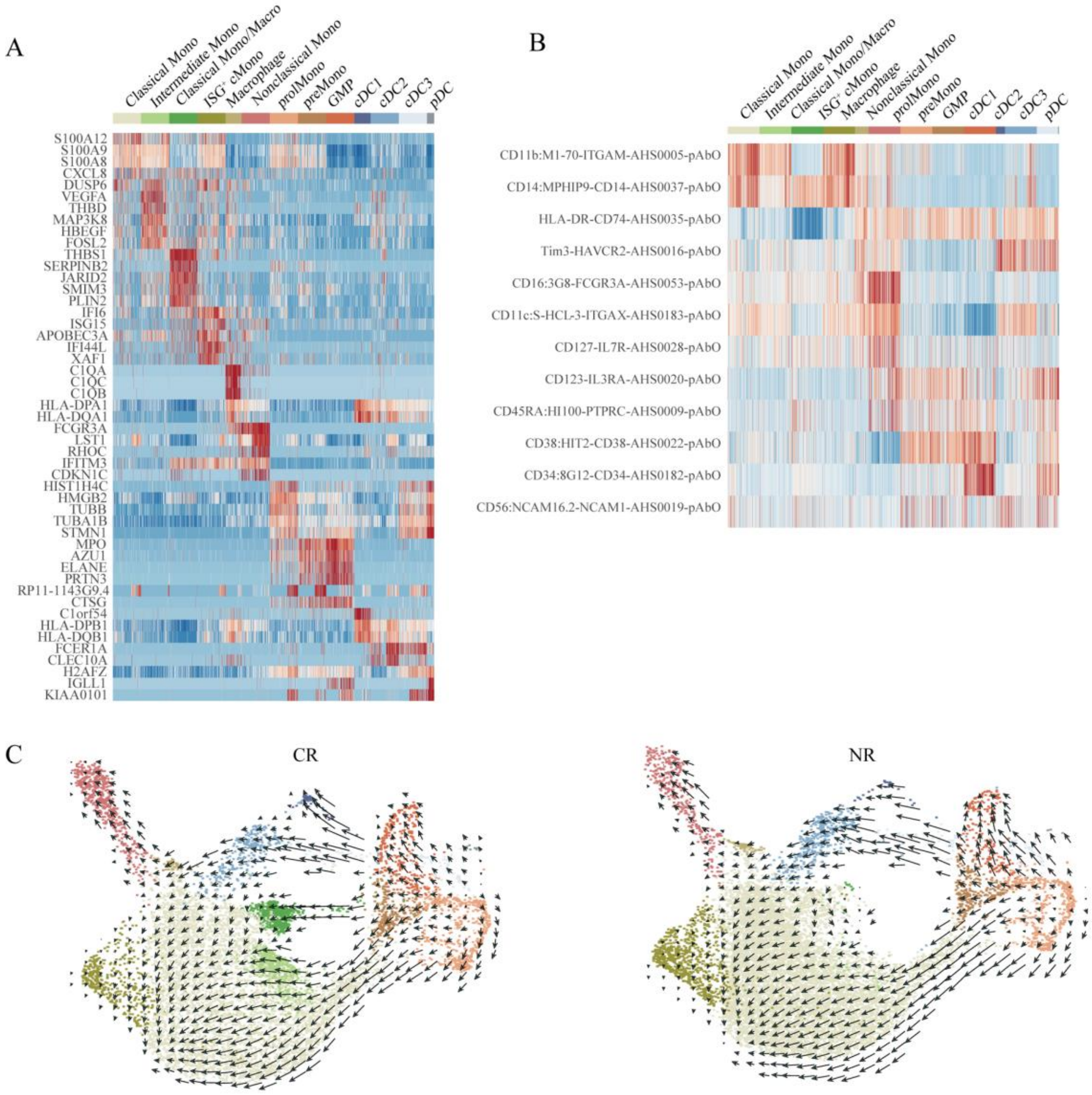


**Supplementary figure 2** Single-cell multimodal omics analysis reveals the composition of myeloid cells in the bone marrow of multiple myeloma patients. (a-b) Heatmaps show the specific high-expression genes (a) and proteins (b) for each myeloid cell subpopulation. (c) Developmental trajectories inferred using scVelo for responder and non-responder groups, with arrow directions indicating the developmental progression.

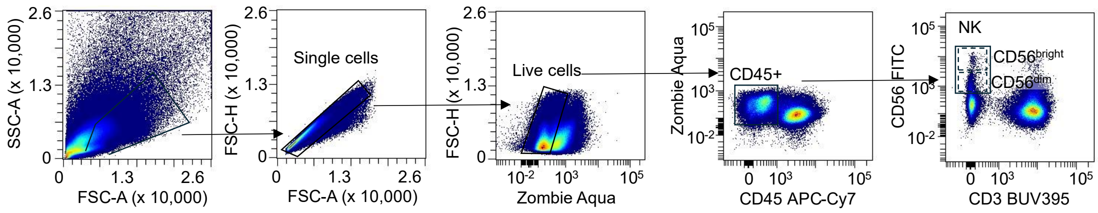


**Supplementary figure 3**. Gating strategy applied for bone marrow aspirate analysis by flow cytometry

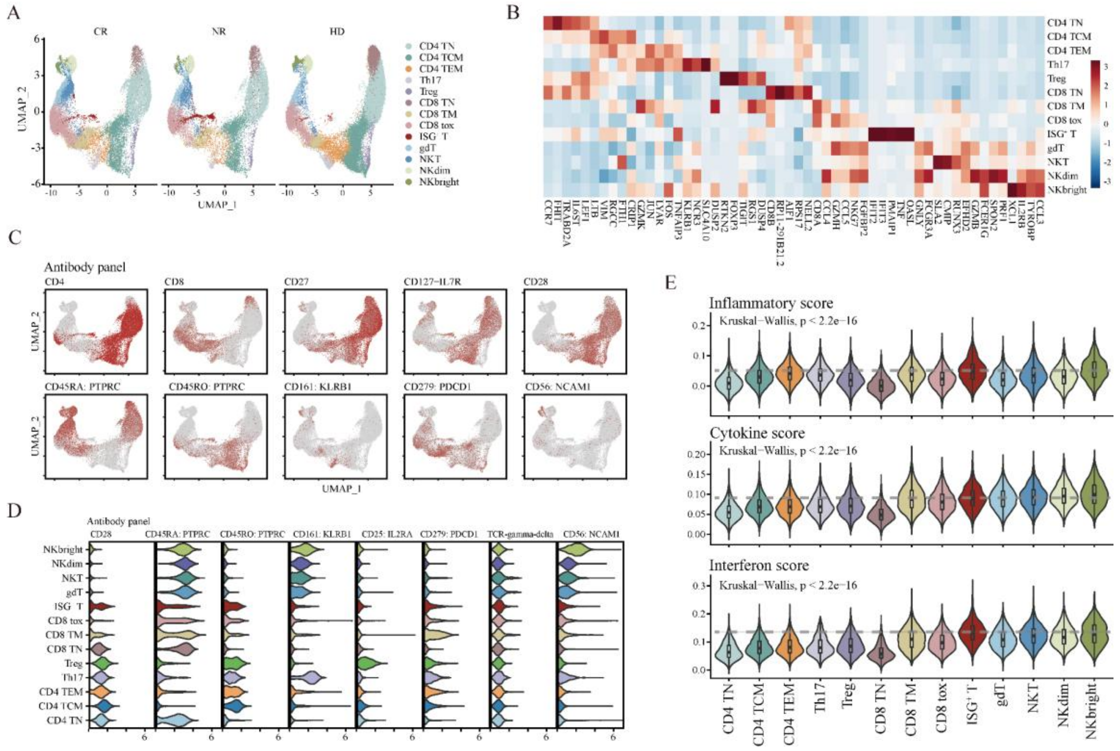


**Supplementary figure 4** (a) UMAP presentation of cell subsets of samples of different phenotypes. (b) Heat maps of gene expression for each subpopulation. (c-d) Display of antibody protein expression levels in each subgroup. (e) Comparison of inflammatory levels, cytokine, and interferon levels scores in different subpopulations.

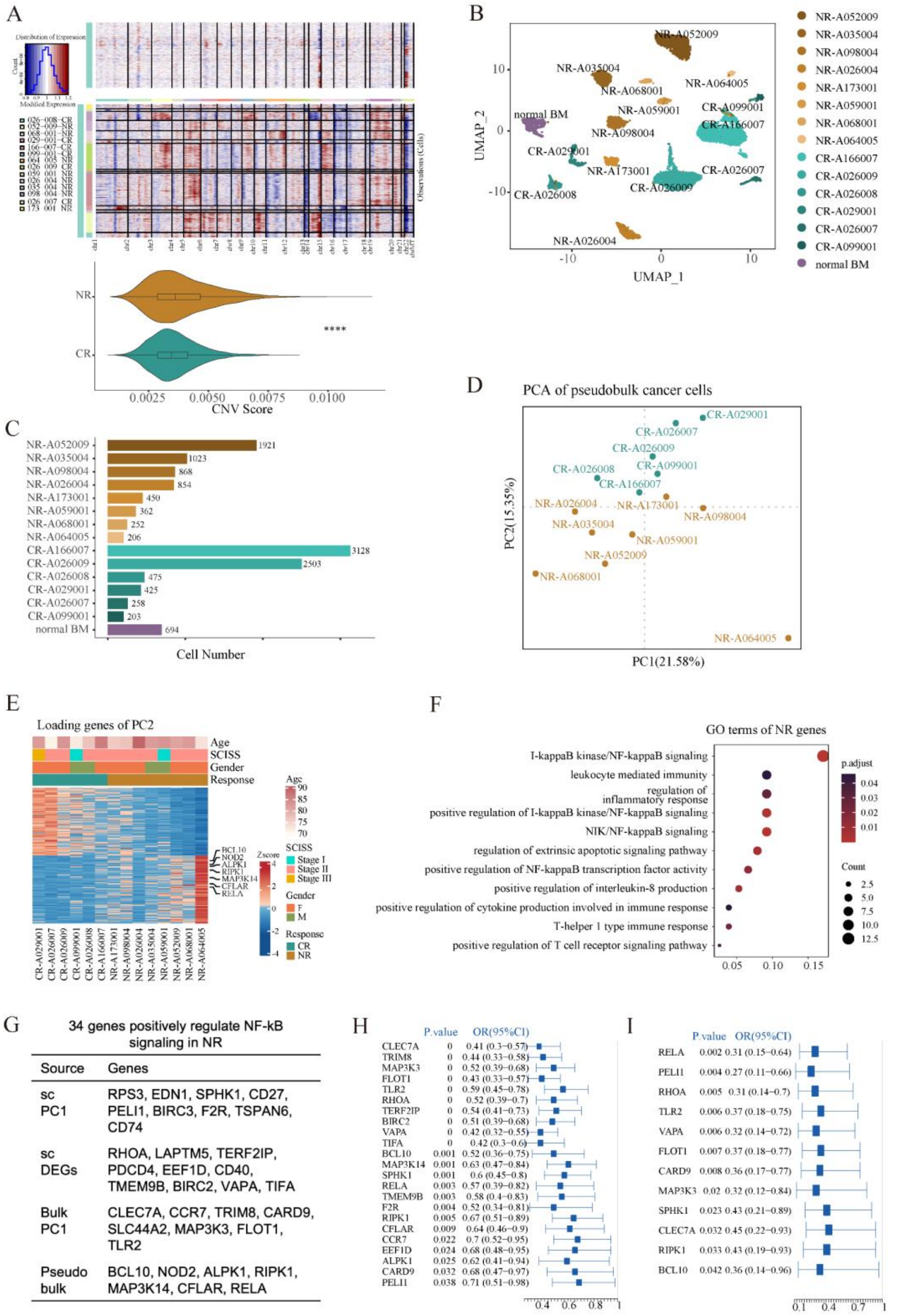


**Supplementary figure 5** Identification and division of cancer cell subpopulations. (a) Heat map of cancer cell CNV scores inferred by InferCNV (top), and box plot of CNV scores in CR and NR groups (bottom). (b) The UMAP shows a fine subpopulation of cancer cells named after the sample. (c) The number of cells corresponding to each fine subpopulation of cancer cells. (d-f) Pseudobulk examined NF-kB enrichment in NR group. (g) Genes positively regulate NF-kB signaling in NR. (h) Poor prognosis of 24 NF-kB genes was verified in 853 patients with multiple myeloma. (i) Poor prognosis of 12 NF-kB genes was verified in 172 Lenalidomide/Daratumumab treated multiple myeloma patients.
